# Socioeconomic inequalities across life and premature mortality from 1971 to 2016: findings from three British birth cohorts born in 1946, 1958, and 1970

**DOI:** 10.1101/2020.04.09.20059410

**Authors:** Meg E Fluharty, Rebecca Hardy, George B. Ploubidis, Benedetta Pongiglione, David Bann

## Abstract

**Introduction:** Disadvantaged socioeconomic position (SEP) in early and adult life has been repeatedly associated with premature mortality. However, it is unclear whether these inequalities differ across time, nor if they are consistent across different SEP indicators.

**Methods:** British birth cohorts born in 1946, 1958 and 1970 were used, and multiple SEP indicators in early and adult life were examined. Deaths were identified via national statistics or notifications. Cox proportional hazard models were used to estimate associations between SEP indicators and mortality risk—from 26-43 (n=40,784), 26-58 (n=35,431), and 26-70 years (n=5,353).

**Results:** More disadvantaged SEP was associated with higher mortality risk—magnitudes of association were similar across cohort and each SEP indicator. For example, hazards ratios (95% CI) between 26-43 years comparing lowest to highest father’s social class were 2.74 (1.02—7.32) in 1946c, 1.66 (1.03—2.69) in 1958c, and 1.94 (1.20—3.15) in 1970c. Childhood social class, adult social class, and housing tenure were each independently associated with mortality risk.

**Conclusions:** Socioeconomic circumstances in early and adult life appear to have had persisting associations with premature mortality from 1971—2016. This reaffirms the need to address socioeconomic factors across life to reduce inequalities in survival to older age.

## Introduction

Previous evidence consistently has consistently shown that disadvantaged socioeconomic position (SEP) in early and adult life is associated with premature mortality risk.[1, 2] However, the magnitude of the inequalities is likely context-specific and may therefore change across time. Evidence on these changes in the UK however, is inconsistent. Inequalities in all-cause mortality by area-level measures of deprivation in adulthood appear to have increased from the 1980s to 2010s in Britain.[3] This contrasts with reports of narrowing inequalities over the same (similar) period by educational attainment[4]—observed trends may therefore be sensitive to the specific SEP indicator used.

Existing studies investigating lifetime SEP and mortality associations have typically been limited to older cohorts (born 1930s-1950s), have used regional, older, or clinical samples; and are limited to single indicators of SEP—with childhood indicators recalled in adulthood.[1, 2] The current study uses three comparable national British birth cohorts—born in 1946, 1958, and 1970—to investigate changes in inequalities in all-cause mortality risk across adulthood and early old age of three generations. The cohorts benefit from multiple prospectively ascertained SEP indicators across life. Given persisting inequalities in multiple diseases and other mortality risk factors across the studied period (1971-2016),[5, 6] we hypothesised that inequalities in premature mortality would have persisted.

## Methods

### Study design and sample

We used data from three British birth cohort studies born in 1946 (MRC National Survey of Health and Development (1946c))[7, 8], 1958 (National Child Development Study (1958c)),[9] and 1970 (British Cohort Study (1970c)).[9] These cohorts have been described in detail elsewhere.[10, 11] As 1946c consists of a social class-stratified sample, analyses were weighted. Participants were included in the current analysis if they had a valid measure of parental and/or own SEP and known vital status and date (from age 26 onwards).

Father’s occupational social class at birth was used in 1958c and 1970c and at age 4 in 1946c (birth data was not used to avoid WWII-related misclassification); occupation was classified using the Registrar General’s Social Class (RGSC) scale: I (professional), II (managerial and technical), IIIN (skilled non-manual), IIIM (skilled manual), IV (partly-skilled), and V (unskilled) occupations. Maternal education collected at birth (1958-1970c) or age 6 (1946c) was distinguished using a binary variable indicating those who continued or left education at the mandatory leaving age.

Own highest educational attainment attained by age 26 (1946c and 1970c) or 23 years (1958c) was categorised into degree/higher, A levels/diploma, O levels/ GCSEs, or none. Participant’s own social class (RGSC), and housing tenure (owner vs renting) were collected at age 26 in 1946c and 1970c and at age 23 in the 1958c.

### Mortality

Death notifications supplied from the Office for National Statistics (ONS), and/or via notifications given to participants’ families during fieldwork. [12, 13]

### Statistical analysis

To aid cross-cohort comparisons, analyses were carried out across the following age ranges: 26-43 years (all cohorts), 26-58 years (1946c-1958c), and 26-70 years (1946c only).

For each SEP measure, cumulative death rates were calculated for each group. Cox proportional hazard models were used to estimate associations between each SEP indicator and all-cause mortality, following checks that the proportional hazard assumption held by calculating Schoenfeld residuals (Supplementary Table S1) and visually inspecting survival curves. Follow-up was from age 26 to date of death or emigration or the end of each follow-up period (age 43, 58 or 70). Follow-up of emigrants and those alive at the end of each period was censored. To provide single quantifications of inequalities, all SEP indicators were converted to ridit scores, resulting in an estimate of the Relative Index of Inequality when included in the models.[14] Cohort differences were formally tested by testing SEP x cohort interaction terms. Models were adjusted for sex. Additionally separate models were run for each sex to investigate whether there were differences in inequalities between men and women. To investigate if associations of SEP across life and premature mortality were cumulative in nature 1) mutually adjusted models were conducted including paternal and own social class—and additionally housing tenure given its suggested importance—with independence of association suggestive of cumulative effects; and 2) a composite lifetime SEP score was used in models, by combining these indicators together and rescaling (with larger effect sizes than each indicator in isolation also suggestive of cumulative effects).[6]

Multiple imputation was conducted to address missing data in SEP indicators (N= 481 (1946c), 514 (1958c), 1,236 (1970c); findings were similar when conducted using complete case analyses. Ten imputed datasets were used and the Nelson-Aalen estimator was included within imputation models. Finally, to investigate if results were similar when examined in the absolute scale, the above models were repeated using logistic regression (dead/alive at the end of each follow-up period with those who emigrated excluded) and estimating the absolute differences in predicted probabilities of mortality in each period. All analyses were conducted in Stata, version 16.0 (StataCorp LP, College Station, Texas).

## Results

In total, 40,784 participants were included in analyses between 26-43 years (13% 1946c, 43% 1958c, 43% 1970c); 35,431 from 26-58 years (23% 1946c, 77% 1946c), and 5,353 from 26-70 years (100% 1946c); see Supplementary Tables S2). Mortality rates were sequentially lower across each cohort— by age 43 they were 74.53 per 1000 person years (1946c), 66.38 (1958c), and 51.63 (1970c); and by age 58 they were 116.94 per 1000 person years (1946c) and 103.66 (1958c).

More disadvantaged SEP was associated with higher mortality risk, with 21 of 24 Hazard Ratios (HRs) being between 1.6 and 3.1 (Figure 1). As anticipated, associations were least precisely estimated at 43 years, where there were fewer deaths; and in the 1946c, which is smaller than 1970c and 1958c. Across each age period, hazard ratios were generally larger in NSHD than the 2 later born cohorts, but the confidence intervals for NSHD at younger ages were wide due to the smaller sample size. (Figure 1 and Supplementary Table S3; all cohort*SEP interaction term p-values were >0.4). For example, HRs of early death from 26–43 years comparing most to least disadvantaged paternal social class were 2.74 (95% CI: 1.02—7.32) in 1946c, 1.66 (1.03—2.69) in 1958c, and 1.94 (1.20— 3.15) in 1970c. Housing tenure was also associated with mortality, renters compared to home owners had a consistently higher risk of death: HRs from 26–43 years were 2.06 (95% CI: 1.03—4.12 in 1946c, 1.30 (0.87—1.94) in 1958c, and 1.61 (1.00—2.60) in 1970c. Associations were weaker for maternal education attainment as an alternative indicator for childhood SEP, particularly for 1946c (Supplementary Table S3).

**Figure 1.**
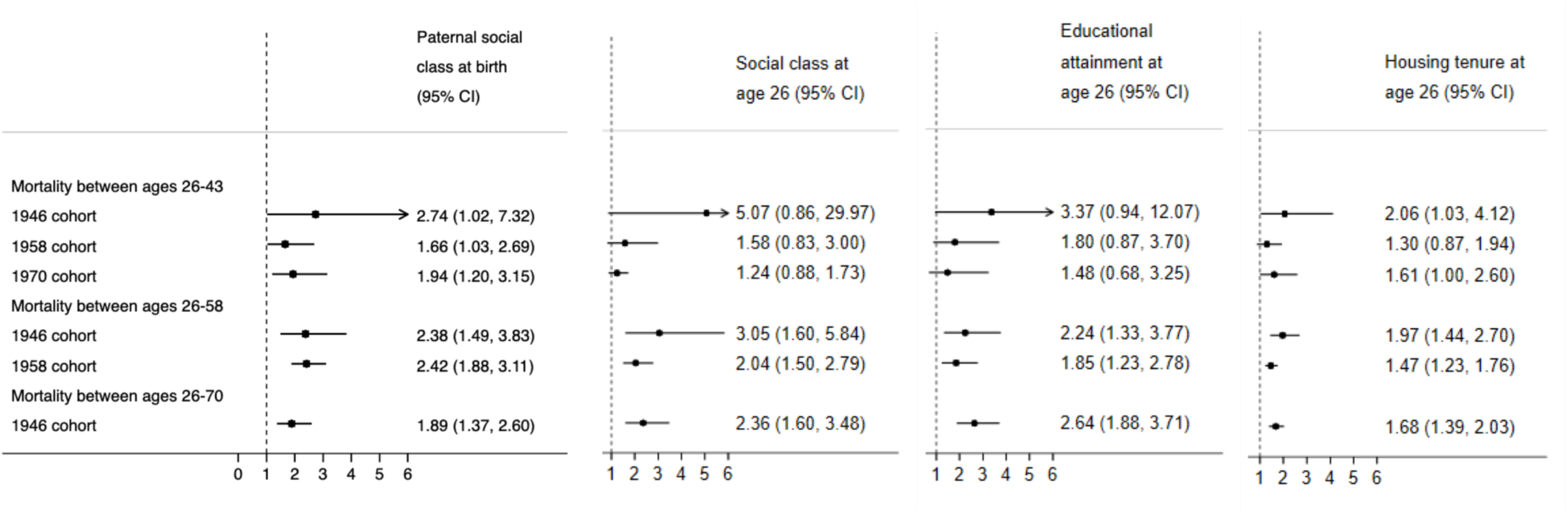
Associations between socioeconomic position in childhood and adult mortality risk: evidence from 3 birth cohort studies *Adjusted for sex* *Cox regression with ridit scores, Hazard Ratios, 95%% CI*

In models including both fathers and own social class, associations were typically partly attenuated but generally still associated with premature mortality. Additionally, there was some evidence that composite lifetime SEP scores had larger magnitudes of association with mortality than each indicator in isolation (particularly in later periods of follow-up; see Supplementary table S4a). Similar findings were found when housing tenure was included in models (Supplementary table S4b).

Findings of persistent inequalities in premature mortality across each cohort were also found when examining on the absolute instead of the relative scale (Supplementary Tables S5), and when conducted separately among men and women (Supplementary Tables S6-7). There was some evidence however for stronger associations among females in the 1946c and among males in the 1970c.

## Discussion

Despite declining mortality rates across the studied period (1971—2016), inequalities in premature mortality appear to have persisted, and were consistently found for multiple SEP indicators in early and adult life. These findings build on prior investigations which have used single birth cohorts[6] or repeated follow-up of adult cohorts.[1, 2]

The persistence of inequalities despite marked changes to population-wide health (such as declines in CVD mortality rates) is suggestive of there being multiple time-depending pathways between SEP and mortality.[15] Further, it is possible that, despite their overlap, each SEP indicator captures independent pathways, resulting in their individual associations with mortality. For example, child SEP is associated with many mortality risk factors such as BMI independently of adult SEP,[10] and housing tenure may specifically capture wealth given the increasing value of housing in Britain— wealth is increasingly suggested to be an important health-relevant SEP indicator. [16] Top causes of death within these cohorts were likely to have been cancers, coronary heart disease, and unnatural causes (i.e., those due to substance abuse and associated liver disease, accidents, assaults and self-injurious behaviour).[17]

This study benefits from the use of three large nationally representative cohort studies, enabling long-run examination of trends in mortality risk, and use of comparable different SEP indicators from childhood and adulthood. While we use multiple indicators of SEP, they are likely to be underestimates of socioeconomic inequality—wealth for example is only crudely approximated by home ownership, we lack comparable data on income across life, and lacked power to investigate social class which may peak in midlife. While there were a small number of participants with missing outcome data, we found the mortality rates in each cohort corresponded with the expected population at the time.[18] Our study was limited to all-cause mortality; however, trends in inequalities may differ by health outcome, e.g. absolute inequalities in coronary heart disease appear to have narrowed in 1994-2008 [19, 20] but inequalities in stroke remained unchanged.[20] As such, future studies with larger sample sizes are warranted to investigate trends in cause-specific premature mortality.

Taken together, our findings re-affirm the need to address socioeconomic factors in both early and adult life in order to reduce inequalities in early-mid adulthood mortality. In contemporaneous and future cohorts, inequalities in premature mortality are likely to be significant barriers to a necessary component of healthy ageing: survival into older age.

## Data Availability

NSHD data are freely accessible to bona fide researchers by applying through the NSHD data sharing website (https://skylark.ucl.ac.uk/NSHD/doku.php).
NCDS and BCS data are freely accessible to bona fide researchers by applying through the UK Data Service (https://www.ukdataservice.ac.uk/).

https://www.ukdataservice.ac.uk/

## Acknowledgements

DB is supported by the Economic and Social Research Council (grant number ES/M001660/1) and MF and DB by The Academy of Medical Sciences / Wellcome Trust (“Springboard Health of the Public in 2040” award: HOP001/1025). RH is Director of CLOSER which is funded by the Economic and Social Research Council (award reference: ES/K000357/1). GBP is Director of Research of CLS which is funded by the Economic and Social Research Council (award reference: ES/M001660/1).The funders had no role in study design, data collection and analysis.

---

**Supplementary Table S1.**
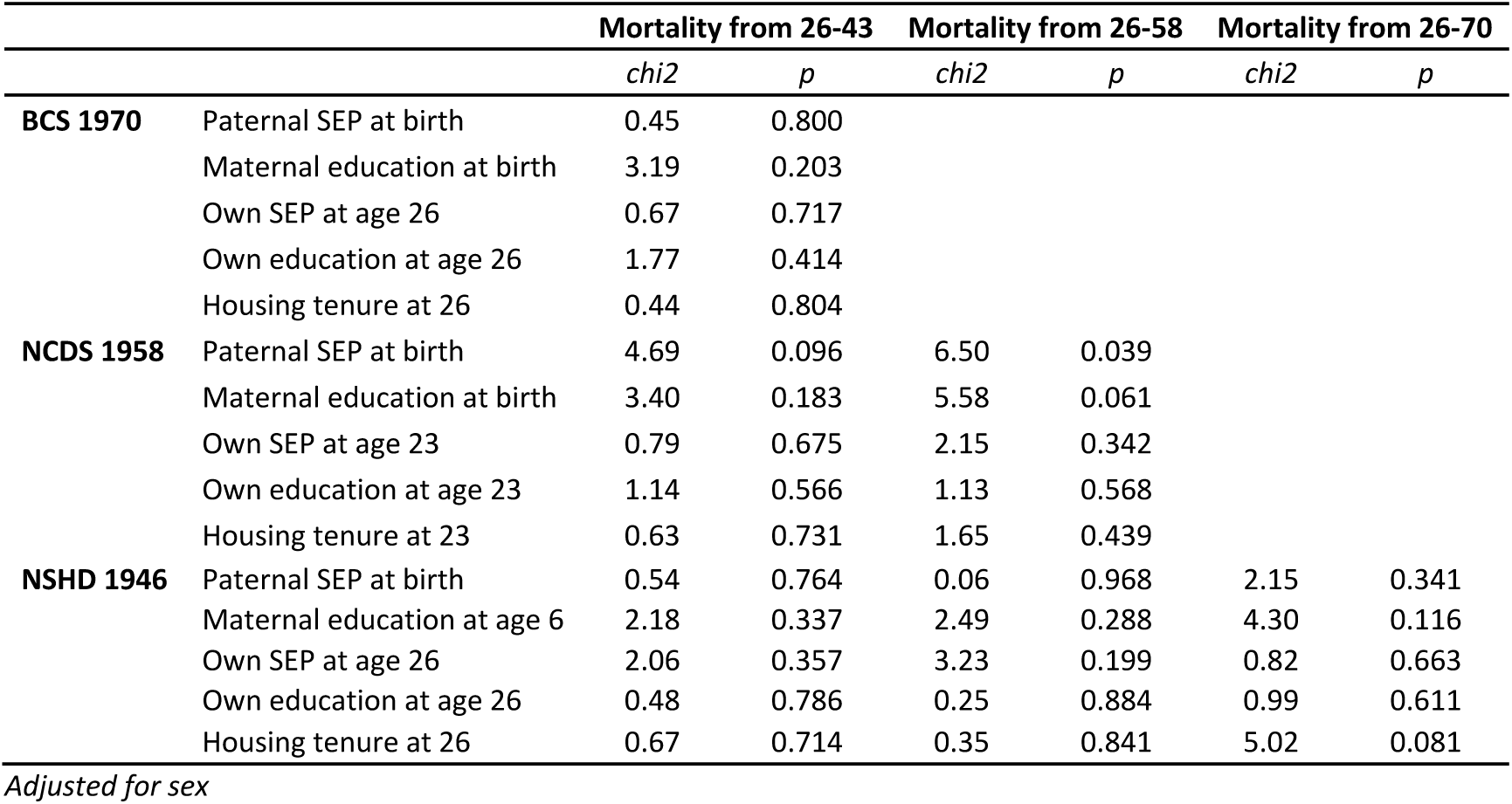
Testing proportional hazards assumption.

|  |  | Mortality from 26-43 |  | Mortality from 26-58 |  | Mortality from 26-70 |  |
| --- | --- | --- | --- | --- | --- | --- | --- |
|  |  | *chi2* | *p* | *chi2* | *p* | *chi2* | *p* |
| **BCS 1970** | Paternal SEP at birth | 0.45 | 0.800 |  |  |  |  |
|  | Maternal education at birth | 3.19 | 0.203 |  |  |  |  |
|  | Own SEP at age 26 | 0.67 | 0.717 |  |  |  |  |
|  | Own education at age 26 | 1.77 | 0.414 |  |  |  |  |
|  | Housing tenure at 26 | 0.44 | 0.804 |  |  |  |  |
| **NCDS 1958** | Paternal SEP at birth | 4.69 | 0.096 | 6.50 | 0.039 |  |  |
|  | Maternal education at birth | 3.40 | 0.183 | 5.58 | 0.061 |  |  |
|  | Own SEP at age 23 | 0.79 | 0.675 | 2.15 | 0.342 |  |  |
|  | Own education at age 23 | 1.14 | 0.566 | 1.13 | 0.568 |  |  |
|  | Housing tenure at 23 | 0.63 | 0.731 | 1.65 | 0.439 |  |  |
| **NSHD 1946** | Paternal SEP at birth | 0.54 | 0.764 | 0.06 | 0.968 | 2.15 | 0.341 |
|  | Maternal education at age 6 | 2.18 | 0.337 | 2.49 | 0.288 | 4.30 | 0.116 |
|  | Own SEP at age 26 | 2.06 | 0.357 | 3.23 | 0.199 | 0.82 | 0.663 |
|  | Own education at age 26 | 0.48 | 0.786 | 0.25 | 0.884 | 0.99 | 0.611 |
|  | Housing tenure at 26 | 0.67 | 0.714 | 0.35 | 0.841 | 5.02 | 0.081 |

*Adjusted for sex*

**Table S2.**
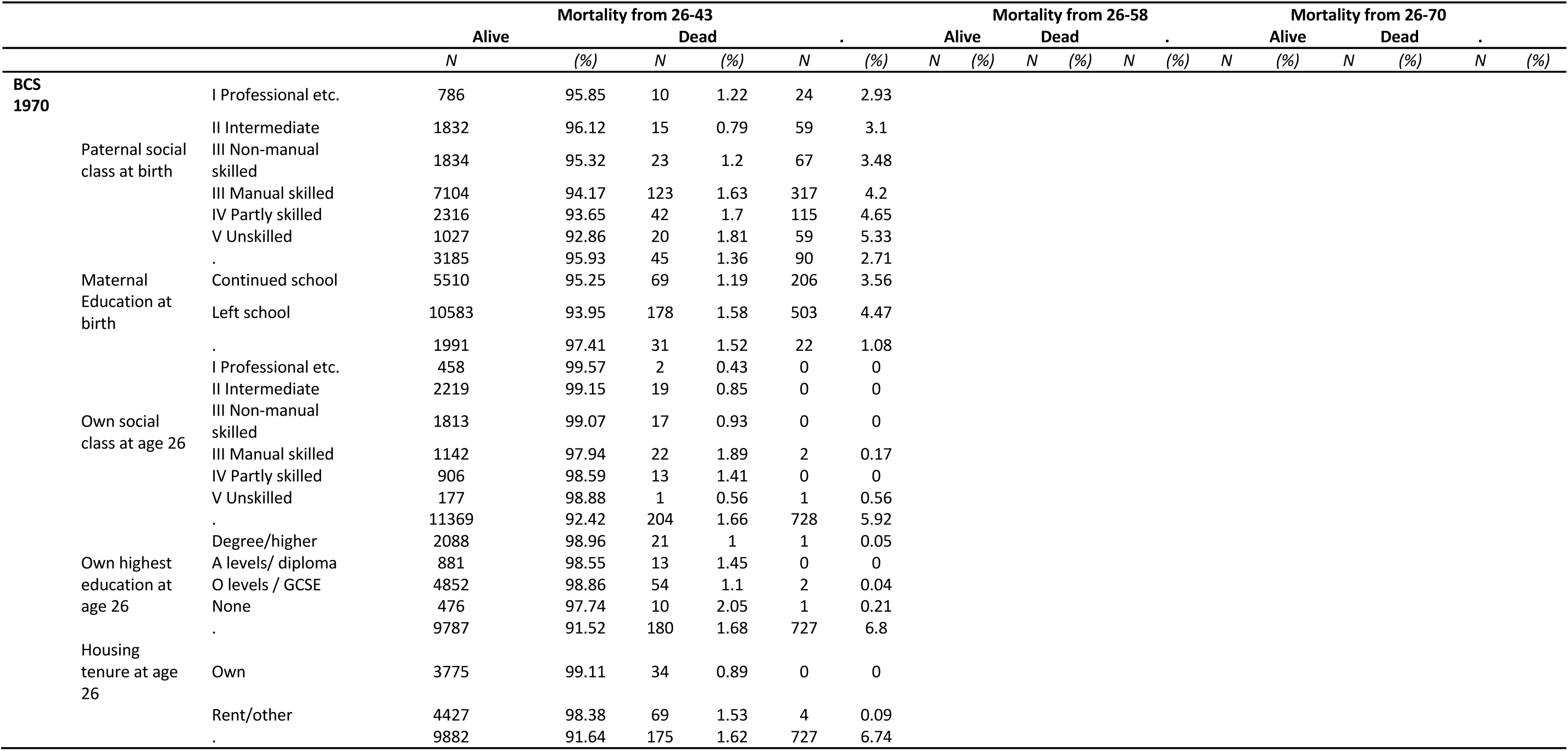
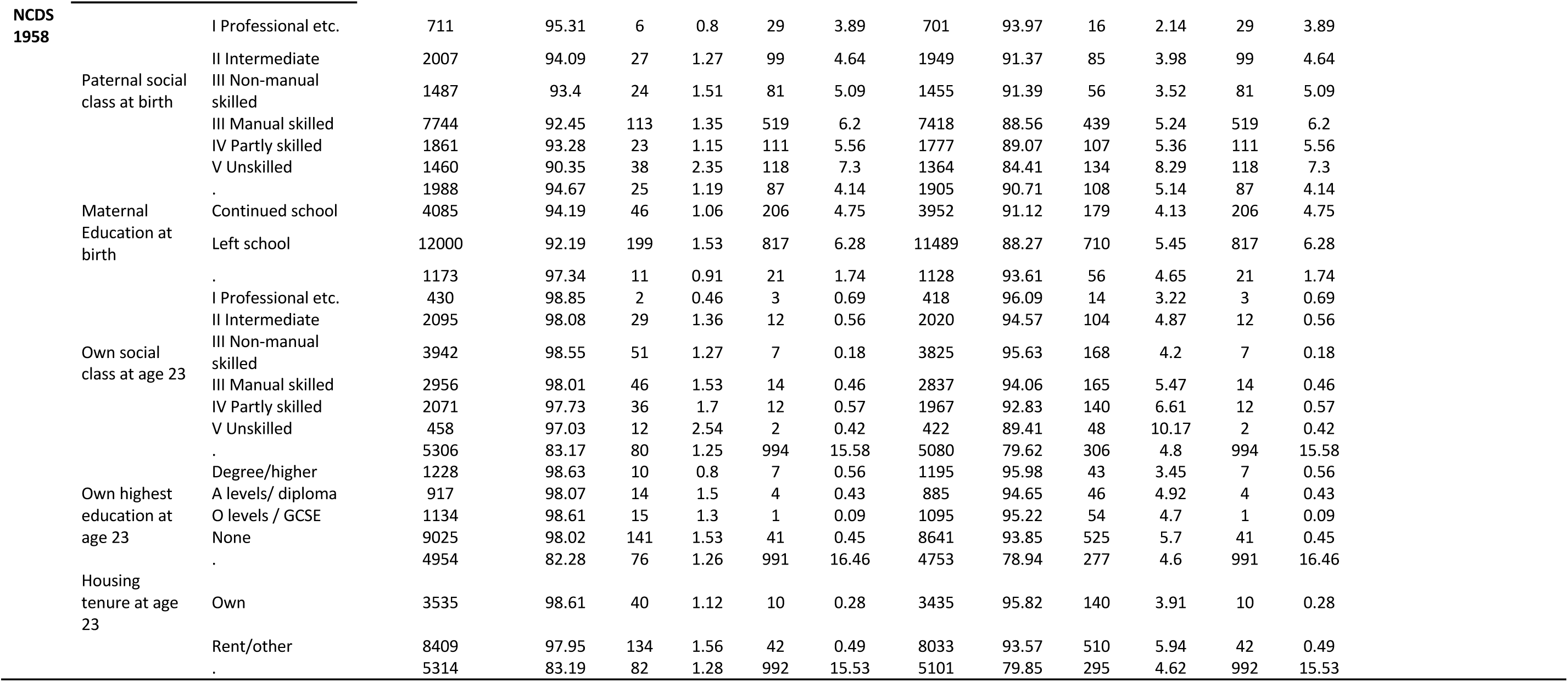
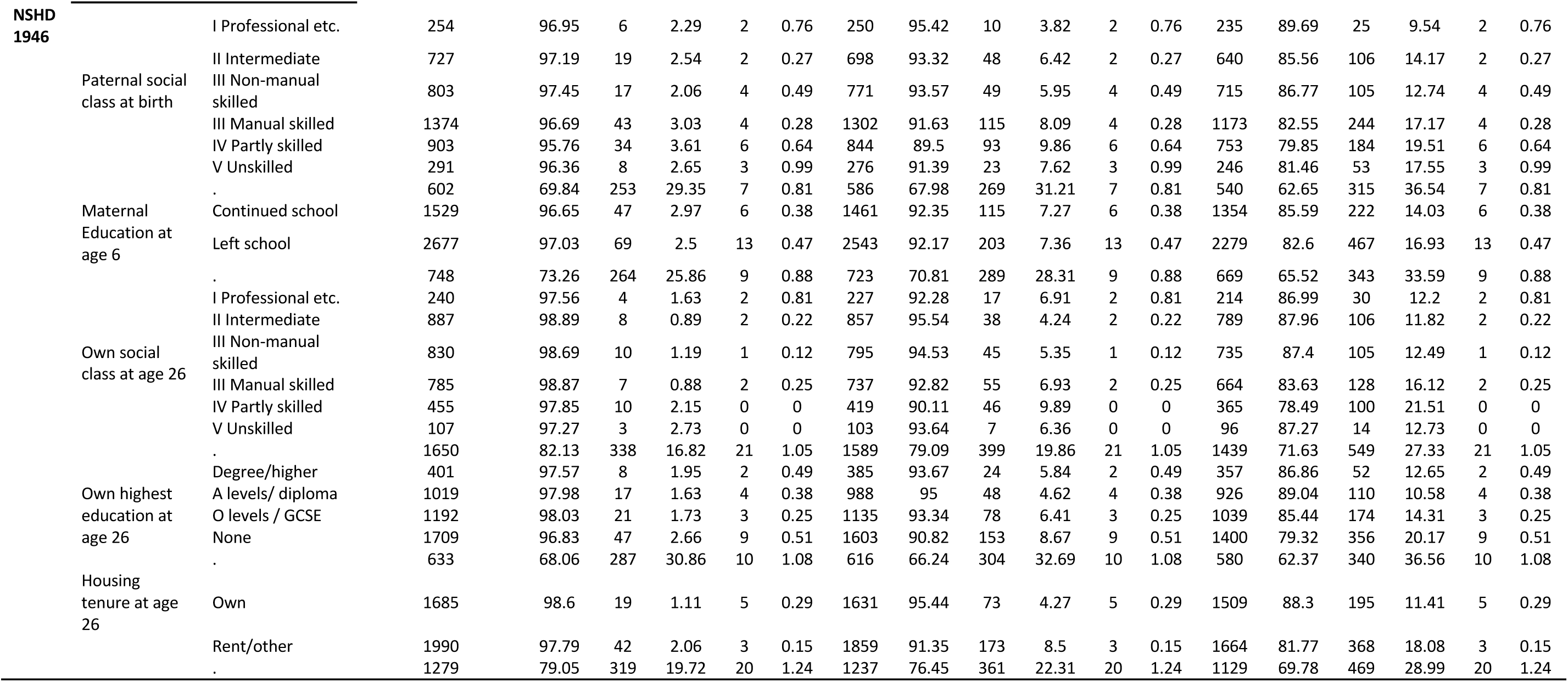
Socioeconomic indicators and mortality status in the 1946, 1958, and 1970 cohort.

**Supplementary Table S3.**
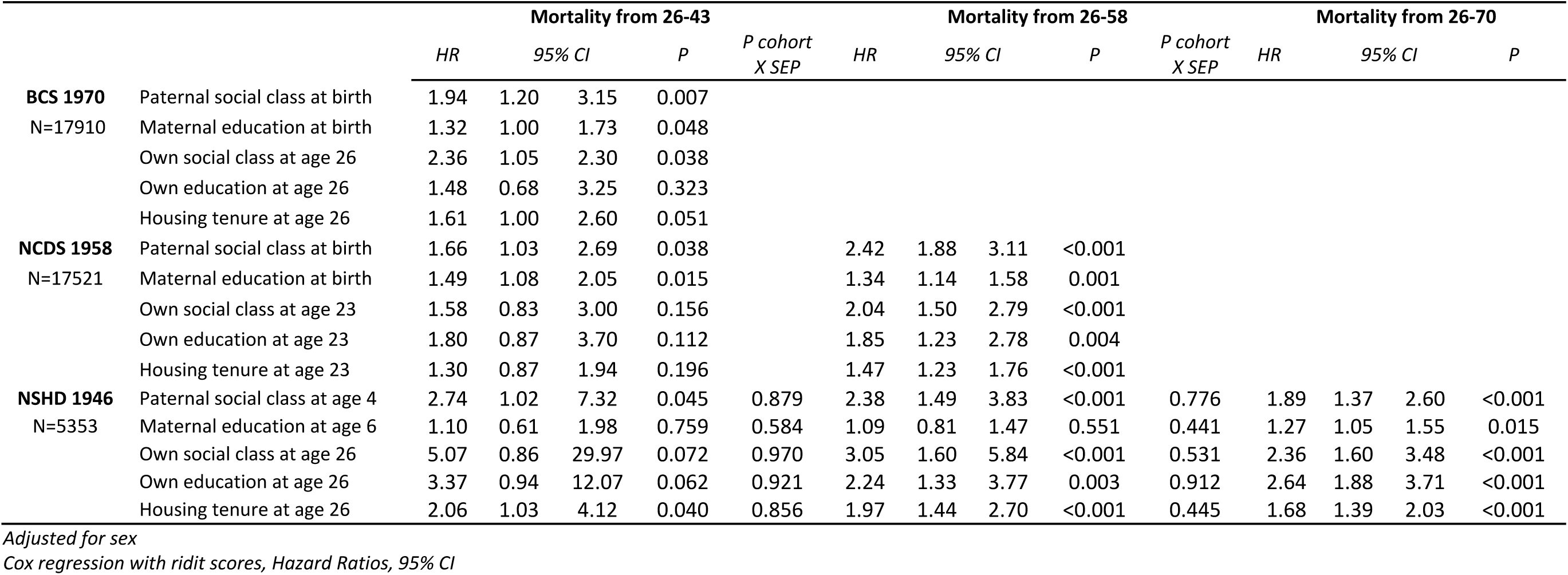
Associations between socioeconomic position in early and adult life and mortality risk: evidence from 3 birth cohort studies, adjusted for sex

**Supplementary Table S4.**
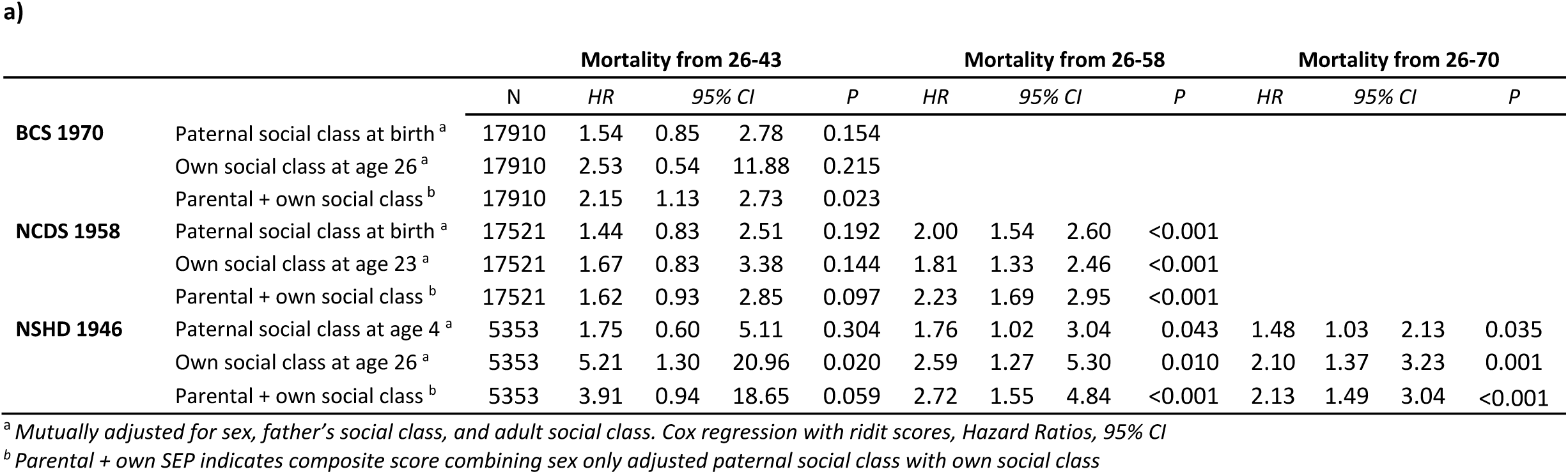
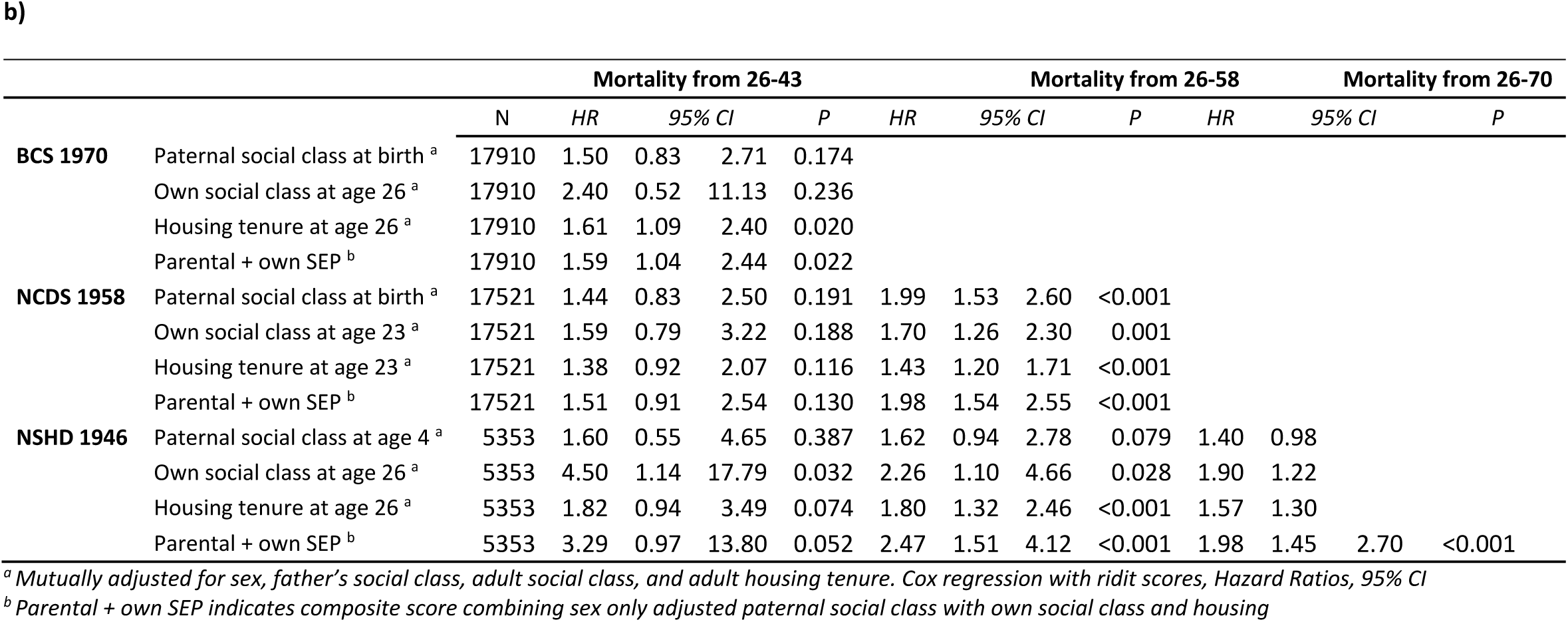
Associations between socioeconomic position in early and adult life and mortality risk: evidence from 3 birth cohort studies, mutually adjusted for multiple indicators of socioeconomic position

**Supplementary Table S5.**
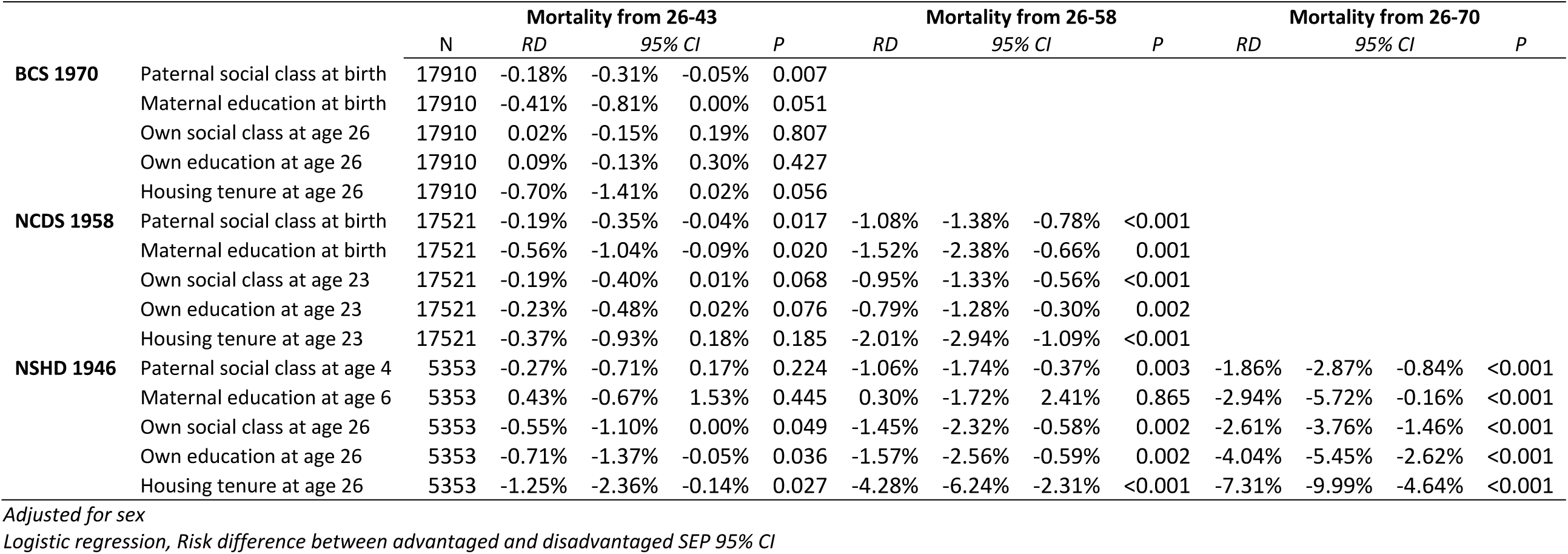
Associations between socioeconomic position in early and adult life and mortality risk: evidence from 3 birth cohort studies, estimated on the absolute scale

|  |  | N | Mortality from 26-43 |  |  |  | Mortality from 26-58 |  |  |  | Mortality from 26-70 |  |  |  |
| --- | --- | --- | --- | --- | --- | --- | --- | --- | --- | --- | --- | --- | --- | --- |
|  |  |  | RD | 95% CI | P | RD | 95% CI | P | RD | 95% CI | P | RD | 95% CI | P |
| **BCS 1970** | Paternal social class at birth | 17910 | -0.18% | -0.31% | -0.05% | 0.007 |  |  |  |  |  |  |  |  |
|  | Maternal education at birth | 17910 | -0.41% | -0.81% | 0.00% | 0.051 |  |  |  |  |  |  |  |  |
|  | Own social class at age 26 | 17910 | 0.02% | -0.15% | 0.19% | 0.807 |  |  |  |  |  |  |  |  |
|  | Own education at age 26 | 17910 | 0.09% | -0.13% | 0.30% | 0.427 |  |  |  |  |  |  |  |  |
|  | Housing tenure at age 26 | 17910 | -0.70% | -1.41% | 0.02% | 0.056 |  |  |  |  |  |  |  |  |
| **NCDS 1958** | Paternal social class at birth | 17521 | -0.19% | -0.35% | -0.04% | 0.017 | -1.08% | -1.38% | -0.78% | <0.001 |  |  |  |  |
|  | Maternal education at birth | 17521 | -0.56% | -1.04% | -0.09% | 0.020 | -1.52% | -2.38% | -0.66% | 0.001 |  |  |  |  |
|  | Own social class at age 23 | 17521 | -0.19% | -0.40% | 0.01% | 0.068 | -0.95% | -1.33% | -0.56% | <0.001 |  |  |  |  |
|  | Own education at age 23 | 17521 | -0.23% | -0.48% | 0.02% | 0.076 | -0.79% | -1.28% | -0.30% | 0.002 |  |  |  |  |
|  | Housing tenure at age 23 | 17521 | -0.37% | -0.93% | 0.18% | 0.185 | -2.01% | -2.94% | -1.09% | <0.001 |  |  |  |  |
| **NSHD 1946** | Paternal social class at age 4 | 5353 | -0.27% | -0.71% | 0.17% | 0.224 | -1.06% | -1.74% | -0.37% | 0.003 | -1.86% | -2.87% | -0.84% | <0.001 |
|  | Maternal education at age 6 | 5353 | 0.43% | -0.67% | 1.53% | 0.445 | 0.30% | -1.72% | 2.41% | 0.865 | -2.94% | -5.72% | -0.16% | <0.001 |
|  | Own social class at age 26 | 5353 | -0.55% | -1.10% | 0.00% | 0.049 | -1.45% | -2.32% | -0.58% | 0.002 | -2.61% | -3.76% | -1.46% | <0.001 |
|  | Own education at age 26 | 5353 | -0.71% | -1.37% | -0.05% | 0.036 | -1.57% | -2.56% | -0.59% | 0.002 | -4.04% | -5.45% | -2.62% | <0.001 |
|  | Housing tenure at age 26 | 5353 | -1.25% | -2.36% | -0.14% | 0.027 | -4.28% | -6.24% | -2.31% | <0.001 | -7.31% | -9.99% | -4.64% | <0.001 |

*Adjusted for sex*
*Logistic regression, Risk difference between advantaged and disadvantaged SEP 95% CI*

**Supplementary Table S6.** Associations between socioeconomic position in early and adult life and mortality risk: evidence from 3 birth cohort studies, stratified by sex

**a)**
|  |  | Males |  |  |  |  |  |  |  |  |  |  |  |  |
| --- | --- | --- | --- | --- | --- | --- | --- | --- | --- | --- | --- | --- | --- | --- |
|  |  | Mortality from 26-43 |  |  |  |  | Mortality from 26-58 |  |  |  | Mortality from 26-70 |  |  |  |
|  |  | N | HR | 95% CI | P |  | HR | 95% CI | P |  | HR | 95% CI | P |  |
| BCS 1970 | Paternal social class at birth | 9239 | 2.45 | 1.39 | 4.30 | 0.002 |  |  |  |  |  |  |  |  |
|  | Maternal education at birth | 9239 | 1.56 | 1.11 | 2.19 | 0.010 |  |  |  |  |  |  |  |  |
|  | Own social class at age 26 | 9239 | 2.52 | 0.65 | 9.77 | 0.166 |  |  |  |  |  |  |  |  |
|  | Own education at age 26 | 9239 | 1.94 | 0.66 | 5.75 | 0.216 |  |  |  |  |  |  |  |  |
|  | Housing tenure at age 26 | 9239 | 1.57 | 0.99 | 2.49 | 0.053 |  |  |  |  |  |  |  |  |
| NCDS 1958 | Paternal social class at birth | 9007 | 1.58 | 0.85 | 2.93 | 0.148 | 2.44 | 1.76 | 3.38 | <0.001 |  |  |  |  |
|  | Maternal education at birth | 9007 | 1.49 | 1.00 | 2.22 | 0.049 | 1.30 | 1.05 | 1.60 | 0.017 |  |  |  |  |
|  | Own social class at age 23 | 9007 | 1.47 | 0.66 | 3.27 | 0.335 | 2.00 | 1.38 | 2.90 | <0.001 |  |  |  |  |
|  | Own education at age 23 | 9007 | 1.96 | 0.74 | 5.17 | 0.172 | 1.71 | 1.03 | 2.83 | 0.037 |  |  |  |  |
|  | Housing tenure at age 23 | 9007 | 1.24 | 0.73 | 2.12 | 0.418 | 1.31 | 1.01 | 1.70 | 0.046 |  |  |  |  |
| NSHD 1946 | Paternal social class at age 4 | 2808 | 2.69 | 0.69 | 10.44 | 0.152 | 2.08 | 1.09 | 3.98 | 0.027 | 1.51 | 0.99 | 2.32 | 0.057 |
|  | Maternal education at age 6 | 2808 | 0.67 | 0.31 | 1.45 | 0.305 | 0.83 | 0.57 | 1.21 | 0.321 | 1.08 | 0.85 | 1.39 | 0.525 |
|  | Own social class at age 26 | 2808 | 5.61 | 0.58 | 53.94 | 0.133 | 3.11 | 1.48 | 6.51 | 0.003 | 2.25 | 1.47 | 3.46 | <0.001 |
|  | Own education at age 26 | 2808 | 4.21 | 0.87 | 20.32 | 0.073 | 1.97 | 1.04 | 3.77 | 0.039 | 2.07 | 1.37 | 3.12 | 0.001 |
|  | Housing tenure at age 26 | 2808 | 2.32 | 0.89 | 6.01 | 0.084 | 1.94 | 1.24 | 3.05 | 0.004 | 1.64 | 1.27 | 2.13 | <0.001 |

*Cox regression with rdit scores, Hazard Ratios, 95% CI* b)
|  |  | Females |  |  |  |  |  |  |  |  |  |  |  |  |
| --- | --- | --- | --- | --- | --- | --- | --- | --- | --- | --- | --- | --- | --- | --- |
|  |  | Mortality from 26-43 |  |  |  |  | Mortality from 26-58 |  |  |  | Mortality from 26-70 |  |  |  |
|  |  | N | HR | 95% CI | P | HR | 95% CI | P | HR | 95% CI | P |  |  |  |
| BCS 1970 | Paternal social class at birth | 8671 | 1.21 | 0.53 | 2.78 | 0.648 |  |  |  |  |  |  |  |  |
|  | Maternal education at birth | 8671 | 1.01 | 0.63 | 1.62 | 0.951 |  |  |  |  |  |  |  |  |
|  | Own social class at age 26 | 8671 | 3.67 | 0.51 | 26.59 | 0.182 |  |  |  |  |  |  |  |  |
|  | Own education at age 26 | 8671 | 1.39 | 0.38 | 5.04 | 0.606 |  |  |  |  |  |  |  |  |
|  | Housing tenure at age 26 | 8671 | 1.91 | 1.13 | 3.23 | 0.016 |  |  |  |  |  |  |  |  |
| NCDS 1958 | Paternal social class at birth | 8514 | 1.84 | 0.83 | 4.07 | 0.135 | 2.21 | 1.47 | 3.31 | <0.001 |  |  |  |  |
|  | Maternal education at birth | 8514 | 1.47 | 0.87 | 2.50 | 0.148 | 1.38 | 1.06 | 1.79 | 0.016 |  |  |  |  |
|  | Own social class at age 23 | 8514 | 1.83 | 0.81 | 4.13 | 0.146 | 1.90 | 1.23 | 2.94 | 0.004 |  |  |  |  |
|  | Own education at age 23 | 8514 | 1.56 | 0.58 | 4.20 | 0.381 | 1.89 | 1.07 | 3.34 | 0.027 |  |  |  |  |
|  | Housing tenure at age 23 | 8514 | 1.39 | 0.86 | 2.24 | 0.181 | 1.54 | 1.20 | 1.98 | 0.001 |  |  |  |  |
| NSHD 1946 | Paternal social class at age 4 | 2545 | 2.80 | 0.62 | 12.65 | 0.181 | 2.84 | 1.45 | 5.55 | 0.002 | 2.65 | 1.66 | 4.24 | <0.001 |
|  | Maternal education at age 6 | 2545 | 2.32 | 0.88 | 6.10 | 0.087 | 1.69 | 1.04 | 2.74 | 0.034 | 1.68 | 1.22 | 2.32 | 0.001 |
|  | Own social class at age 26 | 2545 | 4.64 | 0.50 | 43.41 | 0.172 | 2.99 | 1.06 | 8.47 | 0.040 | 2.53 | 1.27 | 5.07 | 0.010 |
|  | Own education at age 26 | 2545 | 2.53 | 0.33 | 19.28 | 0.364 | 2.78 | 1.13 | 6.83 | 0.026 | 4.45 | 2.46 | 8.06 | <0.001 |
|  | Housing tenure at age 26 | 2545 | 1.86 | 0.73 | 4.77 | 0.193 | 2.01 | 1.25 | 3.24 | 0.004 | 1.73 | 1.29 | 2.34 | <0.001 |

*Cox regression with rdit scores, Hazard Ratios, 95% CI*

**Supplementary Table S7.**
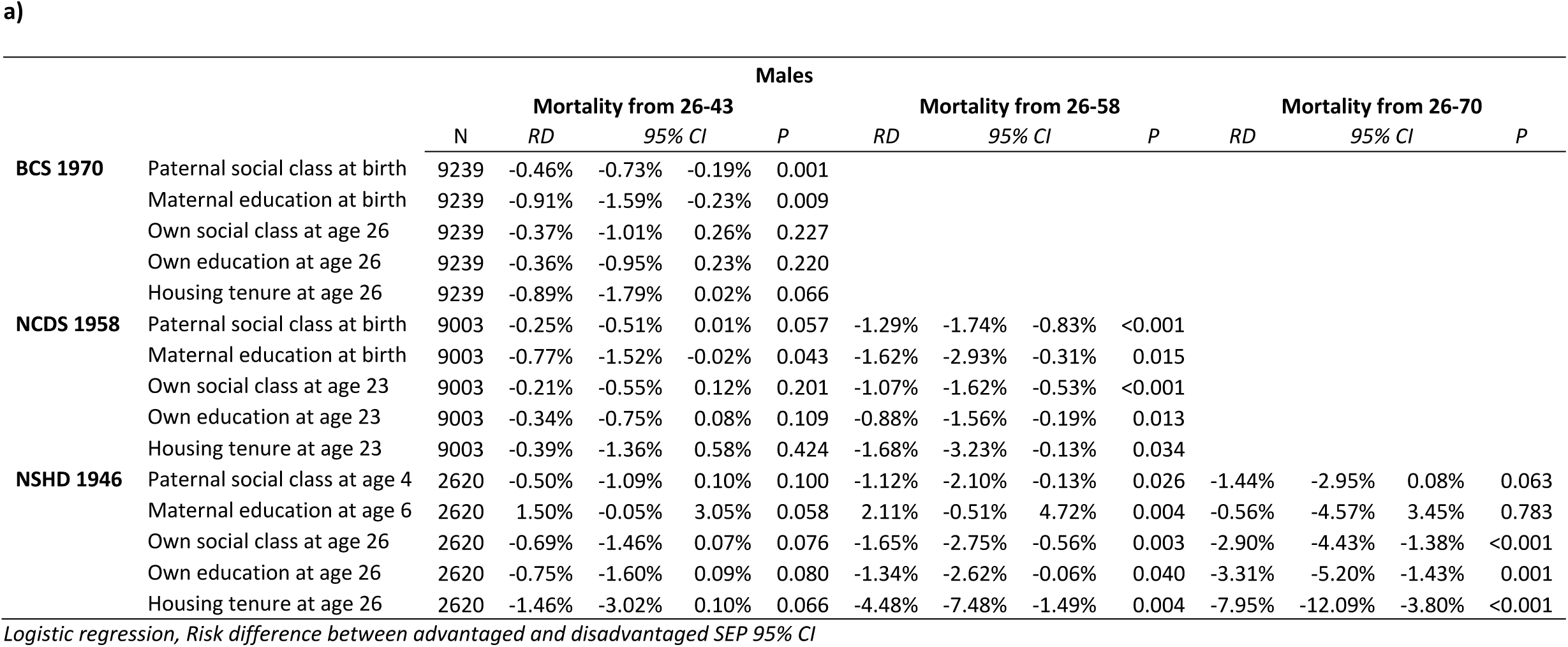
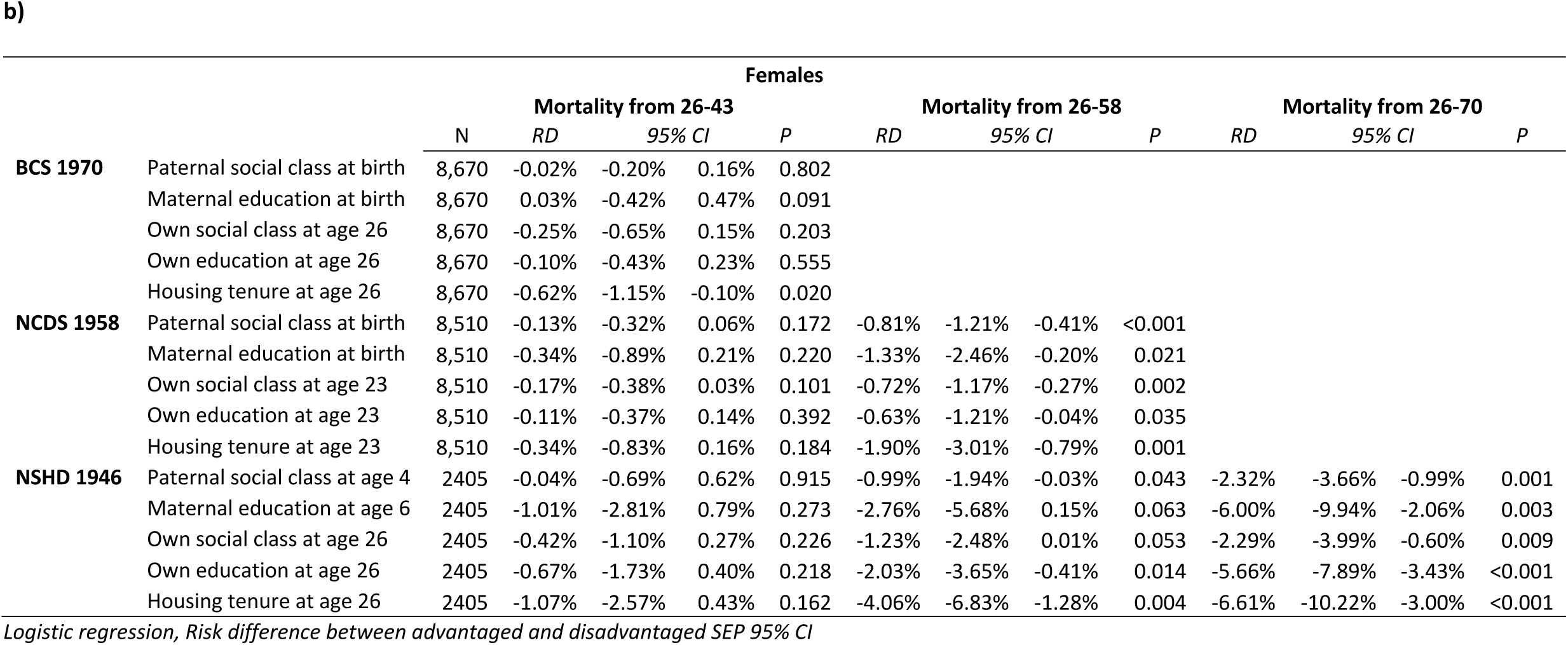
Associations between socioeconomic position in early and adult life and mortality risk: evidence from 3 birth cohort studies, stratified by sex and shown on the absolute scale

## Notes

### Competing Interest Statement

The authors have declared no competing interest.

